# Beyond the pandemic: vaccine hesitancy among youth in rural Malawi as a standing challenge for immunization programming in sub-Saharan Africa - a cross-sectional analytical study

**DOI:** 10.64898/2026.04.30.26352114

**Authors:** Lombani Nyirenda, Ellacy Chirwa, Balwani Chingatichifwe Mbakaya

**Affiliations:** Department of Public Health, University of Livingstonia, Mzuzu, Malawi; School of Medicine, University College Dublin, Dublin, Ireland

**Keywords:** vaccine hesitancy, youth, Malawi, sub-Saharan Africa, immunization programs, pandemic preparedness, COVID-19

## Abstract

**Background:** Vaccine hesitancy among young people in low-resource settings is not a problem that arrived with COVID-19 and departed with it. It is a standing structural challenge that shapes the uptake of every immunization program from routine childhood vaccines to HPV and measles campaigns and that will determine how effectively communities respond to the next epidemic. This study examined COVID-19 vaccine hesitancy among youths aged 18 to 35 years in rural northern Malawi, a population that is systematically under-researched yet critically important for immunization program success. The aim was to generate actionable evidence on the prevalence, demographic predictors, information pathways, and belief-based drivers of hesitancy, applicable to future immunization programming in similar low-resource settings.

**Methods:** A cross-sectional analytical design was employed. Using simple random sampling, 378 participants were recruited from four strategically selected community sites in Nyungwe, Karonga District. Data were collected through structured, pretested, interviewer-administered questionnaires and analysed using IBM SPSS version 26. Descriptive statistics characterised the sample; chi-square tests examined bivariate associations; and binary logistic regression identified independent predictors of hesitancy, adjusting for potential confounders.

**Results:** Vaccine hesitancy was high, affecting 299 of 378 participants (79.1%). Awareness of specific vaccines was moderate: 35% recognised Johnson & Johnson and 34% AstraZeneca, while 31% had never heard of any COVID-19 vaccine. Friends were the dominant information source (40.7%), followed by radio (25.9%), television (24.3%), and WhatsApp (9.0%). Critically, neither vaccine awareness nor information source predicted vaccination status, demonstrating that information exposure alone does not drive uptake. Age was the only statistically significant demographic predictor of hesitancy (OR = 0.60, 95% CI: 0.38–0.94, p = 0.026), with older youths substantially less likely to hesitate. Safety concerns (47%) and perceived personal immunity (30%) were the leading drivers of hesitancy.

**Conclusions:** In rural Malawi, youth vaccine hesitancy is pervasive, age-differentiated, and driven primarily by trust and risk perception deficits rather than information gaps. These findings have immediate relevance for HPV vaccination programs, measles catch-up campaigns, and future epidemic preparedness in sub-Saharan Africa. Immunization strategies that invest in trust-building, peer-mediated communication, and age-targeted risk messaging are more likely to succeed than those relying on information dissemination alone. This study provides evidence that Malawi’s health system and its development partners urgently need to design programs that truly reach and persuade the country’s youth.

## Introduction

### The problem that outlasted the pandemic

On 5 March 2021, Malawi received its first consignment of 360,000 AstraZeneca COVID-19 vaccine doses through the COVAX initiative a moment of hard-won hope in a country whose already fragile health system had been tested to its limits by the pandemic [1]. By October 2021, only 5.8% of eligible young adults aged 18 to 35 years had been vaccinated [2]. In Nyungwe, Karonga District, the site of this study, only 2,250 of an estimated 7,150 eligible young people had accepted a vaccine despite the establishment of new vaccination sites nearby [3]. The vaccines were available. The sites were open. Young people were not coming.

This is not a story that ended when COVID-19 vaccines were rolled back. It is a story about how a generation of young Malawians relates to vaccination itself and that story is still being written. Malawi’s national immunization program continues to fall short of coverage targets for measles, HPV, and other vaccine-preventable diseases, with young people consistently among the hardest cohorts to reach [4]. The World Health Organization identified vaccine hesitancy as one of the ten greatest threats to global health as early as 2019 [5], long before COVID-19 gave the concept global urgency. Understanding why young people in rural Malawi hesitate is therefore not an academic exercise in pandemic retrospection. It is a prerequisite for designing immunization programs that work now and in the future.

### Why young people aged 18 to 35, and why these matter

The deliberate focus of this study on youth aged 18 to 35 years requires justification, because it is a justification with real stakes. This age group is not simply a convenient research population. It is, in the context of sub-Saharan African health systems, simultaneously the most vaccine-hesitant, the most socially connected, and the most consequential demographic for disease transmission and community health outcomes.

Young adults aged 18 to 35 years exhibit what researchers call optimistic bias a well-documented cognitive pattern in which individuals underestimate their personal vulnerability to illness relative to others [6]. They perceive themselves as healthy, resilient, and unlikely to suffer serious consequences from infectious disease. This perception is not irrational; it is a predictable feature of young adult psychology that has been documented across settings from Japan [6] to Portugal [7] and across multiple vaccine-preventable diseases. But in the context of vaccination, optimistic bias is dangerous: it produces the conviction that vaccination is unnecessary precisely among those whose social mobility and interconnectedness makes them key vectors of disease transmission.

In Malawi, this demographic carries additional weight. Young adults aged 18 to 35 constitute a large and growing share of the national population, as in most sub-Saharan African countries [3]. They are the parents of young children who will receive childhood vaccines, the community members who shape local norms about health-seeking behaviour, and the future workforce whose productivity depends on sustained health. They are also the primary target group for HPV vaccination, an intervention with proven cancer-prevention benefits that has faced significant uptake challenges in Malawi and across the region [8]. Understanding and addressing vaccine hesitancy in this group is therefore an investment with returns that extend across generations and across disease areas.

Beyond their demographic importance, young people aged 18 to 35 in Malawi occupy a distinctive information environment. Research consistently shows that peer networks are the dominant source of health information in this group, with friends cited more frequently than health workers, radio, or television as the primary channel through which vaccine-related information is received [9]. This peer-dominated information landscape creates both opportunity and vulnerability: interpersonal networks can carry accurate public health messages efficiently, but they can equally amplify misinformation with remarkable speed, as was documented extensively during the COVID-19 vaccine rollout in Malawi [4] and across sub-Saharan Africa [10].

Despite all of this, youth-specific, community-based vaccine hesitancy data from rural Malawi is almost entirely absent from the published literature. Most existing Malawian studies focus on healthcare workers, mothers, or the general population without disaggregating by age in ways that illuminate youth-specific dynamics [4]. The evidence gap is not simply academic it means that program designers are making decisions about youth immunization communication without the evidence base they need. This study was designed to fill that gap.

### The COVID-19 campaign as a diagnostic tool

This study was conducted within the context of the COVID-19 vaccination campaign, but it should be understood as using that campaign as a diagnostic instrument rather than as its subject. The COVID-19 rollout provided an unusually clear window into vaccine attitudes because it combined high public awareness, significant media attention, widespread community discussion, and a population that had not previously been asked to accept this particular vaccine. It therefore stripped away the habituation effects that can mask hesitancy in routine vaccination programs and revealed the underlying attitudinal landscape with unusual clarity.

What we sought to capture was not whether young Malawians were hesitant about COVID-19 vaccines specifically, but what the structural, social, and demographic factors look like when a rural African youth population is asked to accept a new health intervention under conditions of urgency and imperfect information. Those conditions will recur. The next epidemic, the next new vaccine, the next national immunization campaign will face the same landscape this study maps. The evidence produced here is therefore prospective in its utility even though it is retrospective in its data.

Across sub-Saharan Africa, hesitancy data from the COVID-19 period tells a consistent story: hesitancy is high [11, 12], driven primarily by safety concerns and distrust rather than simple information gaps [9, 13], and shaped by social networks and community belief systems that formal health communication channels have struggled to penetrate [10, 14]. A scoping review of COVID-19 vaccine hesitancy specifically in Malawi found that misinformation was the dominant driver, with community beliefs including associations between vaccines and infertility, disability, and satanic influence playing a significant role [4]. A multi-country study across Burkina Faso, Ethiopia, Ghana, Nigeria, and Tanzania found that perceived lack of vaccine safety and effectiveness were the strongest predictors of hesitancy [12], while research from six African countries demonstrated that trust in government and in society were the most stable and consistent predictors across multiple vaccine types [13]. These are not COVID-specific findings. They are portraits of the immunization challenge in sub-Saharan Africa that will persist long after COVID-19 has receded from public attention.

### Study objectives

This study aimed to: (i) describe the level of COVID-19 vaccine awareness and information sources accessed by youth aged 18 to 35 years in rural northern Malawi; (ii) estimate the prevalence of vaccine hesitancy in this population; (iii) identify independent demographic predictors of hesitancy; and (iv) examine the association between specific beliefs about vaccines and hesitancy. The findings are intended to inform the design of youth-targeted immunization communication strategies in Malawi and comparable low-resource settings in sub-Saharan Africa.

## Materials and methods

### Study design and rationale

A cross-sectional analytical design was employed. This design was selected because it is well-suited to estimating the prevalence of a health-related condition and simultaneously identifying its associations with multiple independent variables within a defined population at a single point in time [15]. Cross-sectional studies are the most widely used design in public health investigations of vaccine hesitancy in low-resource settings [11, 12, 14], enabling comparability of findings across contexts. Although this design does not permit causal inference, it is appropriate for the study objectives, which were descriptive and associational rather than experimental. It is also logistically practicable in resource-limited field settings where longitudinal follow-up is not feasible within the constraints of a community-based study.

### Study setting

The study was conducted in Nyungwe, a rural community in Karonga District, Northern Region of Malawi. Karonga is one of Malawi’s northernmost districts, situated on the shores of Lake Malawi and characterised by a predominantly rural population, smallholder agriculture, and limited access to tertiary health infrastructure. The district’s demographic profile young population, high proportion engaged in subsistence farming and informal trade, low urbanisation, and significant reliance on community-based social networks for health information makes it representative of rural northern Malawi and broadly comparable to other rural low- and middle-income country settings in eastern and southern Africa.

Nyungwe was selected as the study site for substantive rather than merely logistical reasons. At the time of data collection, the community had documented low COVID-19 vaccine uptake among youth despite the establishment of vaccination sites, making it a scientifically appropriate site for investigating the drivers of hesitancy. The community’s characteristics rural, resource-limited, predominantly young, with peer-dominated information networks also represent the type of setting in which immunization programs most urgently need evidence to guide communication strategies.

### Study population and eligibility criteria

The target population comprised all youth aged 18 to 35 years residing in Nyungwe, estimated at 7,150 individuals based on community records and the 2018 Malawi Population and Housing Census [3]. The age range of 18 to 35 years was selected for three reasons that reflect both the scientific literature and Malawi’s programmatic context. First, this range aligns with the WHO definition of youth for health research purposes and corresponds to the priority age cohort identified by Malawi’s Ministry of Health for COVID-19 vaccination [2]. Second, this is the age group in which optimistic bias regarding personal disease vulnerability is most pronounced and in which peer-mediated misinformation is most likely to shape health decisions [6, 7]. Third, this age group represents a critical bridge population in disease transmission dynamics socially mobile, economically active, and highly interconnected making their vaccine acceptance decisions consequential not only for individual protection but for community-level immunity [11]. The scientific justification for targeting this group is therefore not incidental; it reflects a deliberate judgment that understanding hesitancy in this cohort is among the highest-impact investments that vaccine communication research can make.

Participants were eligible if they: (a) were aged 18 to 35 years at the time of data collection; and (b) had resided continuously in the Nyungwe community for at least six months prior to the study, to ensure meaningful exposure to local vaccine communication and community-embedded attitudes. The six-month residency criterion was adopted to exclude transient populations whose attitudes might not reflect the community’s sustained relationship with vaccine messaging. Participants were excluded if they had a serious illness that prevented interview participation or were absent from the community during the data collection period. No eligible participants who were approached declined to participate.

### Sample size determination

Sample size was calculated using Cochran’s formula for cross-sectional prevalence studies [16]:

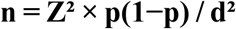

Where Z = 1.96 (corresponding to a 95% confidence interval), p = 0.50 (assumed prevalence of vaccine hesitancy), and d = 0.05 (acceptable margin of error of 5%). The use of p = 0.50 was a deliberate and methodologically conservative choice: in the absence of prior published data on COVID-19 vaccine hesitancy specifically among youth in rural northern Malawi, assuming 50% prevalence maximises the required sample size and therefore minimises the risk of under-powering the study [16]. This approach is consistent with that used in comparable cross-sectional hesitancy studies across sub-Saharan Africa [11, 12]. The formula yielded a minimum required sample of 384 participants. Although a 10% non-response adjustment was built into the planning (yielding an adjusted target of approximately 422), all 378 sampled participants completed the survey, achieving a 100% response rate. This exceeded the minimum required sample, and all planned analyses were adequately powered.

### Sampling strategy

Participants were selected using simple random sampling from a sampling frame constructed from available community records, including school registers, Vision Club membership lists, and village registers compiled in collaboration with local community leaders. Simple random sampling was chosen over convenience or purposive approaches because it provides every eligible individual in the sampling frame with an equal probability of selection, thereby minimising selection bias and producing a statistically representative sample [17]. This is particularly important in a study examining the determinants of hesitancy, where a non-random sample could systematically over- or under-represent particular attitudinal groups.

To ensure that the diversity of youth experience within Nyungwe was adequately represented, the sample was stratified by recruitment site. Four sites were included, each capturing a distinct subgroup of young people: Nyungwe Secondary School (in-school youth, representing youth who are embedded in formal educational institutions and exposed to structured health information); Vision Club (youth in organised community groups, representing those with higher civic engagement); Ryangawuwo Village (household-based rural youth, representing those most embedded in traditional community networks and most likely to rely on peer information channels); and Nyungwe Market (youth employed in the informal economy, representing economically active youth outside formal education). This multi-site stratification was not merely pragmatic it was a deliberate design decision to prevent the common research error of conflating the attitudes of students with those of the broader youth population, and to ensure that findings reflect the genuine heterogeneity of rural youth life circumstances. Proportional allocation across sites ensured that each subgroup contributed to the final sample in proportion to its estimated share of the local youth population.

### Data collection instrument

Primary data were collected using a structured, interviewer-administered questionnaire. This mode was selected for three reasons rooted in the characteristics of the study population and setting. First, interviewer administration ensures standardisation of question delivery across participants, allows trained data collectors to clarify ambiguities in real time, and reduces measurement error attributable to misunderstood questions [18]. Second, it is appropriate for populations with heterogeneous literacy levels as is the case in rural Malawi, where a proportion of participants had attained only primary education because it does not require participants to read or write independently. Third, face-to-face interviewer administration consistently achieves higher response rates and more complete data than self-administered instruments in community settings in low- and middle-income countries [18].

The questionnaire was originally developed in English by the research team, drawing on validated instruments used in comparable regional studies [11, 12]. It was subsequently translated into Chichewa and Tumbuka the two most widely spoken local languages in Karonga District using a rigorous forward-translation and back-translation process to ensure linguistic accuracy and conceptual equivalence across language versions. The instrument covered four thematic domains: (i) sociodemographic characteristics, including age, sex, highest education level, occupation, household size, and distance to the nearest health centre; (ii) COVID-19 vaccine awareness and sources of vaccine-related information; (iii) vaccination status and self-reported hesitancy; and (iv) specific beliefs and attitudes about COVID-19 vaccines, including perceived safety, efficacy, personal immunity, and supernatural associations. Vaccine hesitancy was operationally defined as a delay in accepting or a refusal of COVID-19 vaccination despite the availability of vaccination services, consistent with the WHO SAGE working group definition [5].

The questionnaire was pretested on 30 youth residing in a community outside the study area to assess clarity, cultural appropriateness, logical flow, and average completion time. Items identified as ambiguous, culturally inappropriate, or consistently misunderstood were revised before final deployment. Internal consistency of the full instrument was assessed using Cronbach’s alpha, which yielded a coefficient of 0.82, indicating good reliability [19]. Each interview lasted approximately 20 to 30 minutes and was conducted in a private space at the relevant recruitment site to protect participant confidentiality and reduce social desirability bias. Data collection took place between May and July 2023.

### Variables

The primary outcome variable was COVID-19 vaccine hesitancy, operationalised as a binary measure (hesitant vs. not hesitant) based on participants’ self-reported delay in accepting or refusal of COVID-19 vaccination. Independent variables included: age (continuous, and grouped as 18–22, 23–27, or 28–35 years); sex (male/female); highest education level (primary, secondary, or tertiary); occupation (student, farmer, business, or working class); household size (1–3, 4–6, or >6 members); distance to the nearest health centre (<1 km, 1–5 km, >5 km); primary source of vaccine information (friends, radio, television, or WhatsApp); and specific beliefs about COVID-19 vaccines (fear of serious reactions; belief that the vaccine is a hoax; belief that it remains under trial; belief in high personal immunity; belief that the vaccine is unsafe; belief that vaccination leads to death; and belief that vaccines are associated with Satanism). Socioeconomic status, religious affiliation, and prior health-seeking behaviour were considered as potential confounders and controlled for in multivariable models.

### Statistical analysis

All data were entered, cleaned, and analysed using IBM SPSS Statistics, version 26. Data cleaning involved systematic checking for completeness, logical consistency, and out-of-range values before analysis commenced. The item non-response rate was less than 5% across all variables; missing values were excluded listwise from analyses involving the affected items, and complete-case analysis was used throughout.

Descriptive statistics characterised the study sample. Continuous variables are reported as means and standard deviations; categorical variables as frequencies and valid percentages. Chi-square (χ²) tests were used to assess bivariate associations between categorical variables specifically, between vaccine awareness and primary information source, and between information source and vaccination status. Chi-square testing was appropriate for these analyses because all variables were categorical and cell expected frequencies exceeded five in more than 80% of cells [20].

Binary logistic regression was used to identify independent predictors of vaccine hesitancy. Two separate models were estimated: the first included sociodemographic predictors (sex, age, education, occupation, and household size); the second examined belief-based predictors (the seven attitudinal items listed above). Binary logistic regression was selected over alternatives because the outcome variable is dichotomous, and this method estimates the probability of the outcome occurring as a function of multiple predictor variables simultaneously, controlling for confounding [21]. Adjusted odds ratios (AOR) with 95% confidence intervals (CI) are reported for all modelled associations. The significance threshold was set at p < 0.05 for all tests. Model fit was assessed using the Hosmer-Lemeshow goodness-of-fit test. Age was analysed both as a continuous variable and in categorical form; the categorical approach was retained in the final model because it accommodates non-linear age effects and produces estimates that are more directly interpretable in a programmatic context. Stratified analyses by sex and education level explored potential effect modification. Sensitivity analyses using alternative categorisations of age and education were conducted to test the robustness of results to classification decisions.

### Ethical considerations

Ethical approval was granted by the University of Livingstonia Research Ethics Committee (reference: UNILIA-REC/UGS/64/2023). The study was conducted in full accordance with the ethical principles of the Declaration of Helsinki. Prior to data collection, all participants received a participant information sheet in their preferred language (English, Chichewa, or Tumbuka), explaining the study’s purpose, procedures, risks, benefits, confidentiality protections, voluntary nature of participation, and their right to withdraw at any time without consequence. Written informed consent was obtained from all participants before any data were collected. All questionnaires were assigned unique numeric identifiers; no names or other directly identifying information were recorded in the dataset. Data were stored securely, with access restricted to the named members of the research team.

## Results

### Sociodemographic characteristics of participants

A total of 378 participants completed the survey, achieving a 100% response rate. The sample was approximately balanced by sex: 194 males (51.3%) and 184 females (48.7%). Most participants were in the two younger age groups: 146 (38.6%) were aged 18–22 years and 133 (35.2%) were aged 23–27 years, with 99 (26.2%) aged 28–35 years. The majority had completed secondary education (n = 241, 63.8%), followed by tertiary (n = 110, 29.1%) and primary (n = 27, 7.1%). Students constituted the largest occupational group (n = 268, 70.9%), reflecting the inclusion of Nyungwe Secondary School as a recruitment site, followed by farmers (n = 70, 18.5%) and those in business (n = 36, 9.5%). Nearly half of all participants (n = 182, 48.1%) lived in households of four to six members, and the majority (n = 229, 60.6%) resided within one kilometre of a health centre. All participants were from rural settings. Full sociodemographic details are presented in Table 1.

**Table 1.** Sociodemographic characteristics of study participants (n = 378).

| Characteristic | n | % |
| --- | --- | --- |
| Sex |  |  |
| Male | 194 | 51.3 |
| Female | 184 | 48.7 |
| Age group (years) |  |  |
| 18–22 | 146 | 38.6 |
| 23–27 | 133 | 35.2 |
| 28–35 | 99 | 26.2 |
| Highest education level |  |  |
| Primary | 27 | 7.1 |
| Secondary | 241 | 63.8 |
| Tertiary | 110 | 29.1 |
| Occupation |  |  |
| Student | 268 | 70.9 |
| Farmer | 70 | 18.5 |
| Business | 36 | 9.5 |
| Working class | 4 | 1.1 |
| Household size (members) |  |  |
| 1–3 | 93 | 24.6 |
| 4–6 | 182 | 48.1 |
| >6 | 103 | 27.2 |
| Distance to nearest health centre |  |  |
| <1 km | 229 | 60.6 |
| 1–5 km | 132 | 34.9 |
| >5 km | 17 | 4.5 |

### Vaccine awareness and information sources

Awareness of specific COVID-19 vaccines was moderate in this sample. The most widely recognised vaccine was Johnson & Johnson, cited by 134 participants (35.4%), closely followed by AstraZeneca, cited by 128 (33.9%). Notably, 116 participants (30.7%) reported that they had never heard of any COVID-19 vaccine a substantial proportion given that these individuals lived within one kilometre of a health centre and within a community that had been the subject of active vaccination outreach. This level of awareness is considerably lower than the 76.1% overall vaccine knowledge reported among pilgrims in Saudi Arabia [22] and reflects significant information access inequalities between rural African communities and populations with greater exposure to formal health media.

Friends were the most commonly cited source of vaccine-related information (n = 154, 40.7%), followed by radio (n = 98, 25.9%), television (n = 92, 24.3%), and WhatsApp (n = 34, 9.0%). The dominance of peer networks as the primary information channel is consistent with patterns documented across sub-Saharan Africa [9, 11] and underscores both the opportunity and the risk embedded in interpersonal communication: peers can carry accurate health messages efficiently, but they are equally effective at transmitting misinformation [4, 10].

A chi-square test revealed a statistically significant association between the vaccine a participant had heard of most and their primary information source (χ² = 12.706, df = 6, p = 0.048; Pearson’s r = 0.158, p = 0.002), indicating that different information channels shaped familiarity with specific vaccines differently. However, and this is a finding of central programmatic importance information source was not significantly associated with vaccination status (χ²(3) = 0.326, p = 0.955). Participants who received their vaccine information from friends were no more likely to have been vaccinated than those who relied on radio, television, or WhatsApp. Awareness, regardless of its source, did not translate into action. These findings are presented in Table 2.

**Table 2.** Vaccine awareness by primary information source, and vaccination status by information source.

| Vaccine heard of most | Friends n (%) | Radio n (%) | TV n (%) | WhatsApp n (%) | Total n |
| --- | --- | --- | --- | --- | --- |
| AstraZeneca | 61 (47.7) | 35 (27.3) | 24 (18.8) | 8 (6.3) | 128 |
| Johnson & Johnson | 59 (44.0) | 28 (20.9) | 35 (26.1) | 12 (9.0) | 134 |
| Never heard of any | 34 (29.3) | 35 (30.2) | 33 (28.4) | 14 (12.1) | 116 |
| $\chi^2 = 12.706$, $df = 6$, $p = 0.048$; Pearson's $r = 0.158$, $p = 0.002$; Spearman's $\rho = 0.160$, $p = 0.002$ | | | | | |
| Vaccination status | Friends n (%) | Radio n (%) | TV n (%) | WhatsApp n (%) |  |
| Vaccinated | 19 (12.3) | 11 (11.2) | 12 (13.0) | 5 (14.7) |  |
| Not vaccinated | 135 (87.7) | 87 (88.8) | 80 (87.0) | 29 (85.3) |  |
| $\chi^2(3) = 0.326$, $p = 0.955$ no significant association between information source and vaccination status. | | | | | |

### Prevalence of vaccine hesitancy

Vaccine hesitancy was highly prevalent: 299 of 378 participants (79.1%) reported hesitancy toward COVID-19 vaccination, while only 79 (20.9%) reported no hesitancy. This figure substantially exceeds hesitancy rates reported in several comparable settings across sub-Saharan Africa. Wollburg and colleagues documented hesitancy ranging from 5% to 37% across six East and West African countries [14], while Abubakari and colleagues found hesitancy rates of 29% to 65% across five African countries [12]. The 79% rate observed here may partly reflect the specific characteristics of this population: rural, young, predominantly peer-informed, and embedded in a community context characterised by active circulation of vaccine misinformation, as documented in the Malawian literature [4]. The gap between 5.8% vaccination coverage nationally among this age group [2] and 79% hesitancy in this sample is itself a finding of programmatic significance it suggests that hesitancy, not merely access, is a primary driver of low uptake in this cohort.

### Demographic predictors of vaccine hesitancy

Binary logistic regression examining demographic predictors of hesitancy found that age was the only statistically significant predictor (B = −0.514, Wald = 4.967, p = 0.026; OR = 0.60, 95% CI: 0.38–0.94). Each step upward in age category was associated with approximately 40% lower odds of hesitancy, meaning that participants in the 28–35 years group were substantially less likely to hesitate than those aged 18–22 years. Sex, education, occupation, and household size were not statistically significant predictors of hesitancy in this sample, though education showed a non-significant trend toward higher hesitancy among more educated participants (OR = 2.20, p = 0.081). Full regression results are presented in Table 3.

**Table 3.** Binary logistic regression: demographic predictors of vaccine hesitancy (n = 378).

| Variable | B | S.E. | Wald | df | p | OR (95% CI) |
| --- | --- | --- | --- | --- | --- | --- |
| Sex | -0.437 | 0.261 | 2.798 | 1 | 0.094 | 0.65 (0.39–1.08) |
| Age<sup>1*</sup> | -0.514 | 0.231 | 4.967 | 1 | 0.026 | 0.60 (0.38–0.94) |
| Education | 0.789 | 0.452 | 3.051 | 1 | 0.081 | 2.20 (0.91–5.33) |
| Occupation | 0.499 | 0.363 | 1.891 | 1 | 0.169 | 1.65 (0.81–3.36) |
| Household size | 0.066 | 0.183 | 0.130 | 1 | 0.718 | 1.07 (0.74–1.54) |
| Constant | -3.450 | 2.196 | 2.467 | 1 | 0.116 | 0.03 |

\* $p < 0.05$ . <sup>1</sup>Age entered as ordinal category (18–22 = 1; 23–27 = 2; 28–35 = 3). CI = confidence interval; OR = odds ratio; S.E. = standard error.

### Belief-based drivers of vaccine hesitancy

The most commonly cited reason for hesitancy was the perception that COVID-19 vaccines are unsafe, reported by 178 participants (47.1%). This was followed by belief in sufficiently high personal immunity to make vaccination unnecessary (n = 114, 30.2%), fear of death post-vaccination (n = 49, 13.0%), belief that vaccines carry a supernatural or satanic association (n = 23, 6.1%), fear of serious side effects (n = 8, 2.1%), belief that vaccines are a hoax (n = 4, 1.1%), and belief that vaccines remain under clinical trial (n = 2, 0.5%).

The safety concern finding mirrors what has been reported across sub-Saharan Africa. Wang and colleagues found that perceived lack of safety was the strongest predictor of hesitancy among adolescents across five African countries (aPR: 3.52, 95% CI: 3.00–4.13) [11]. Abubakari and colleagues similarly found that uncertainty about vaccine safety was among the most common reasons for hesitancy among adults across the region [12]. In Malawi specifically, the scoping review by Nkambule and Mbakaya identified safety misinformation including beliefs that vaccines cause infertility, disability, or death as a primary driver of hesitancy [4]. The prominence of perceived high immunity as a second major driver reflects the optimistic bias documented among young adults in multiple settings [6, 7], and is particularly concerning because it is rooted in a cognitive misperception rather than a factual misunderstanding that can be corrected through information alone.

Logistic regression examining the independent association of specific beliefs with hesitancy produced results that are counterintuitive and scientifically important. None of the beliefs typically associated with hesitancy in higher-income settings showed statistically significant associations in this sample: fear of serious reactions (OR = 1.93, p = 0.298), belief that vaccines are a hoax (OR = 0.62, p = 0.666), belief that vaccines remain under trial (OR = 1.56, p = 0.599), belief in high personal immunity (OR = 0.96, p = 0.862), belief that vaccines are unsafe (OR = 0.99, p = 0.987), and belief that vaccination leads to death (OR = 0.81, p = 0.597) were all non-significant. The single significant predictor was belief that vaccines are associated with Satanism, which was paradoxically associated with substantially lower odds of hesitancy (OR = 0.27, 95% CI: 0.17–0.43, p < 0.001). Table 4 presents the full regression results for belief-based predictors.

**Table 4.** Binary logistic regression: belief-based predictors of vaccine hesitancy.

| Belief | B | S.E. | Wald | df | p | OR (95% CI) |
| --- | --- | --- | --- | --- | --- | --- |
| Fear of serious reactions | 0.655 | 0.630 | 1.082 | 1 | 0.298 | 1.93 (0.56–6.63) |
| Vaccine is a hoax | −0.472 | 1.094 | 0.186 | 1 | 0.666 | 0.62 (0.07–5.37) |
| Vaccine still under trial | 0.448 | 0.852 | 0.277 | 1 | 0.599 | 1.57 (0.29–8.39) |
| High personal immunity | −0.046 | 0.265 | 0.030 | 1 | 0.862 | 0.96 (0.57–1.61) |
| Vaccine is not safe | −0.005 | 0.287 | 0.000 | 1 | 0.987 | 0.99 (0.57–1.74) |
| Vaccination leads to death | −0.208 | 0.394 | 0.280 | 1 | 0.597 | 0.81 (0.37–1.76) |
| Vaccine linked to Satanism* | −1.300 | 0.238 | 29.754 | 1 | <0.001 | 0.27 (0.17–0.43) |

\* $p < 0.001$ . CI = confidence interval; OR = odds ratio; S.E. = standard error.

## Discussion

### Principal findings and their significance

This study set out to understand why young people in rural northern Malawi hesitate to accept vaccines using COVID-19 as the instrument of inquiry but asking a question that extends far beyond it. What we found is both clear and sobering. Vaccine hesitancy is not a marginal phenomenon in this population. It is the dominant response, affecting nearly four in every five young people surveyed. It is not primarily a product of information gaps; it is rooted in safety fears, misperceptions of personal immunity, and a relationship with official health institutions that does not yet generate sufficient trust to overcome those fears. And it is age-differentiated: younger people within the 18-to-35 cohort are meaningfully more hesitant than their older peers. Each of these findings has direct implications for how immunization programs in Malawi and across sub-Saharan Africa should be designed.

### The information paradox: why knowing is not enough

One of the most programmatically consequential findings of this study is that neither vaccine awareness nor information source predicted vaccination status. A participant who had heard of AstraZeneca or Johnson & Johnson was no more likely to have been vaccinated than one who had never heard of any COVID-19 vaccine. A participant who received vaccine information from a friend was no more likely to have been vaccinated than one who relied on radio, television, or WhatsApp. Information, regardless of its content or channel, did not produce action.

This finding challenges the logic of the information-deficit model the still-prevalent assumption that hesitancy is primarily a product of insufficient knowledge and that providing accurate information to the public will naturally translate into behaviour change [23]. The evidence from this study joins a growing body of literature demonstrating that this model is inadequate. Osuagwu and colleagues found across sub-Saharan Africa that social media users, TV viewers, and those who relied on friends and family for vaccine information showed higher rates of vaccine resistance despite being well-informed [10]. Wollburg and colleagues demonstrated that vaccine acceptance levels in sub-Saharan Africa remained far above vaccination rates, implying that attitudinal barriers are not the only obstacle access and delivery barriers operate alongside hesitancy even among those who express willingness to vaccinate [14].

The deeper mechanism is trust. When individuals do not trust the source of information, the vaccine itself, or the institutions recommending it, information is filtered through a lens of suspicion that neutralises its persuasive effect. Unfried and Priebe demonstrated, across three vaccine types and six African countries, that trust in government and trust in society were the most consistent and stable predictors of hesitancy more predictive, and more stable across contexts, than any other variable examined [13]. This is a crucial insight for program design. Investing in communication volume more messages, more channels, more frequency without first addressing the trust deficit is unlikely to achieve meaningful improvements in uptake.

### Age as the primary demographic lever

Age was the only statistically significant demographic predictor of hesitancy in this study, with each step toward an older age group associated with approximately 40% lower odds of hesitancy. This pattern is consistent with published evidence. Soares and colleagues, in a study of vaccine hesitancy in Portugal, identified younger age as one of the strongest predictors of hesitancy [7], while Wang and colleagues found comparable age gradients among adolescents and young adults across five sub-Saharan African countries [11]. The consistency of this finding across such diverse settings from southern Europe to rural Africa suggests that it reflects something fundamental about the psychology of young adulthood rather than context-specific factors.

The most plausible explanation is optimistic bias: the well-documented tendency of younger adults to perceive themselves as less vulnerable to serious illness than others, and to discount the personal relevance of health interventions accordingly [6]. Young people in this sample were not ignoring vaccination out of ignorance. Many of them were aware of the vaccine. They simply did not believe they needed it. This is a qualitatively different challenge from the one that more information can solve it requires engagement with risk perception at a personal and community level, through trusted messengers who can speak credibly about why vaccination matters even for the healthy and young.

The trend toward higher hesitancy among more educated participants while not statistically significant is consistent with patterns observed in Malawi’s broader population and across sub-Saharan Africa [14]. Wollburg and colleagues found that hesitancy was higher in richer, better-educated households and in urban areas [14]. One interpretation is that more educated youth have greater exposure to and perhaps greater engagement with vaccine-related content on social media platforms such as WhatsApp, where misinformation circulates widely and persuasively [4, 10]. This finding complicates the assumption that education is a reliable protective factor against hesitancy and suggests that digital media literacy may be as important as formal education in shaping vaccine attitudes among this generation.

### Safety concerns and perceived immunity: the real barriers

The dominance of safety concerns (47%) and perceived immunity (30%) as drivers of hesitancy in this sample reflects patterns that have been documented consistently across sub-Saharan Africa and that are not specific to COVID-19 vaccines. Wang and colleagues found that perceived lack of safety was the strongest predictor of hesitancy across five African countries (aPR: 3.52) [11], while Abubakari and colleagues found that concerns about safety and effectiveness were the most commonly endorsed reasons for hesitancy across the region [12]. Evidence from the Democratic Republic of Congo shows that actual post-vaccination adverse events are predominantly mild and transient [24], yet the perception that vaccines are dangerous persists robustly in communities where this misinformation has circulated and has not been effectively rebutted.

The gap between perceived and actual risk is the central challenge for safety communication. Factual correction alone presenting data on adverse event profiles is insufficient when safety concerns are embedded in social networks as community-held beliefs rather than individual misconceptions. What is needed is community-grounded engagement that validates the concern, provides credible and locally relevant evidence, and comes from messengers who have earned trust through consistent and honest engagement over time. Faith leaders, community health workers, and respected peer figures have been identified as the most effective channels for this kind of communication in sub-Saharan African settings [11, 13]. Formal health authorities, acting alone, are unlikely to shift these beliefs through information campaigns.

The prominence of perceived immunity as a hesitancy driver has particular implications for future outbreak response. When a new vaccine is introduced for a disease that young people perceive as not affecting them seriously as was the case with COVID-19 among youth the perceived immunity argument becomes a powerful barrier. Immunization communication must engage directly with this perception, providing evidence that natural immunity is not reliable or permanent, and that vaccination offers protection that individual health status alone cannot guarantee.

### The Satanism finding: contextual complexity and methodological humility

The finding that belief in a satanic association with vaccines was significantly associated with lower odds of hesitancy (OR = 0.27, p < 0.001) is the most counterintuitive result in this study and deserves honest, contextualised interpretation. In published studies from Europe and North America, conspiracy theories and supernatural beliefs are consistently associated with greater hesitancy [23, 25]. The opposite direction observed here is not simply a statistical anomaly.

One plausible interpretation draws on Malawi’s religious landscape. The Satanism narrative also documented in the Malawian scoping review [4] generated substantial community discussion, including in church settings where faith leaders actively rebutted the association between vaccines and satanic influence. In communities where trusted religious leaders publicly affirmed vaccine safety in response to this narrative, it is possible that those who heard and internalised the satanic claim were also exposed to the most direct and credible rebuttals and that the process of community engagement around this belief ultimately produced greater vaccine acceptance among those who engaged with it most deeply. An alternative explanation is statistical: the Satanism belief was reported by only 6.1% of the sample, producing small cell sizes that may yield unstable logistic regression estimates.

What this finding illustrates, regardless of its mechanism, is that the relationship between beliefs and behaviour is context-dependent and cannot be assumed to operate uniformly across settings. Quantitative survey instruments are powerful tools for estimating prevalence and identifying associations, but they cannot capture the social processes through which beliefs circulate, are contested, and are ultimately resolved in community life. Future research should complement quantitative approaches with qualitative inquiry focus groups, in-depth interviews, ethnographic observation to understand the meaning and social trajectory of specific vaccine beliefs in this and similar communities. This is not merely a methodological recommendation; it is a prerequisite for designing communication strategies that are actually responsive to the communities they seek to serve.

### What this means for Malawi and for sub-Saharan Africa

Malawi is a country at a health crossroads. It is a signatory to global immunization targets that require sustained youth engagement, it is implementing HPV vaccination programs for adolescent girls, and it is rebuilding trust in health systems after a pandemic that in Malawi as elsewhere generated both new investment in immunization infrastructure and new waves of community suspicion about vaccine intentions. The evidence produced by this study speaks directly to this moment.

The findings support four specific programmatic priorities. First, peer-based communication must be resourced as a primary strategy, not a supplementary one. Since friends are the dominant information channel for young people in this community, training and supporting community health ambassadors drawn from youth’s own social networks young people who can speak with credibility to their peers about vaccine safety, personal risk, and community responsibility is likely to achieve far greater impact than mass media campaigns or health worker outreach alone. Wollburg and colleagues identified the creation of positive social norms around vaccination, leveraging trusted and accessible sources, as among the most promising policy options for boosting uptake in sub-Saharan Africa [14].

Second, age-differentiated strategies are needed within the youth cohort itself. The finding that 18-to-22-year-olds are substantially more hesitant than 28-to-35-year-olds despite both groups being targeted under the same age-range definition suggests that a single communication approach applied across the 18-to-35 group will systematically underserve its youngest and most hesitant members. Younger youth may require more intensive engagement, including peer-led small-group sessions that can address optimistic bias directly and personally.

Third, trust must be treated as a program input, not a program output. Trust is not something that emerges automatically when good vaccines are delivered efficiently. It must be built through sustained, honest, two-way engagement with communities engagement that precedes campaigns, continues during them, and does not disappear between them. Health systems in Malawi and across sub-Saharan Africa that invest in this kind of relationship-based community engagement will have a structural advantage in every future immunization effort.

Fourth, this study’s findings are not relevant only to Malawi. The structural conditions it documents rural youth with peer-dominated information environments, safety fears rooted in misinformation, trust deficits, and age-related optimistic bias are present across sub-Saharan Africa and indeed across many LMIC settings globally. The evidence generated here contributes to a regional and global evidence base that program designers, funders, and policymakers can draw on for routine immunization programs, epidemic preparedness planning, and health system strengthening investment decisions.

### Limitations

Several limitations of this study must be acknowledged. First, the cross-sectional design does not permit causal inference; we can describe associations at a single point in time but cannot establish the direction of relationships or rule out unmeasured confounding. Second, the study was conducted in a single rural community in northern Malawi, which limits generalisability to other geographic, demographic, or socioeconomic contexts. Third, self-reported data are susceptible to social desirability bias; participants may have under-reported hesitancy or over-reported vaccine knowledge in the interview context, potentially leading to underestimation of the true prevalence of hesitancy. Fourth, the sampling frame was constructed from community records that may incompletely capture highly mobile or socially marginalised youth subgroups. Fifth, the quantitative design does not allow for in-depth exploration of the reasoning, lived experiences, or cultural contexts that underpin vaccine attitudes, limiting the interpretive reach of the findings. Future research combining quantitative and qualitative approaches, and engaging with longitudinal designs where feasible, would substantially strengthen the evidence base in this area.

## Conclusions

This study was motivated by a question that goes beyond COVID-19: what does the immunization landscape look like for young people in rural Malawi, and what does it tell us about what needs to change? The answer is both sobering and actionable. Vaccine hesitancy is pervasive affecting four in five young people and is rooted not in ignorance but in a deficit of trust and a misperception of personal risk that information campaigns alone cannot address. Age is the most important demographic predictor, with younger youth being substantially more hesitant and therefore requiring targeted, age-appropriate, peer-mediated engagement. Safety concerns and perceived immunity are the dominant drivers, and they call for community-grounded, trust-building communication rather than mass information delivery.

The 378 young people who participated in this study told us, in their own words and through their responses, why they would not accept a vaccine that was available to them. They told us that they were worried it was not safe. They told us they thought their own bodies would protect them. They told us they heard about it from friends. They did not tell us they lacked information they told us they lacked confidence. Translating that message into program design is the work that now needs to happen, in Malawi, and across the sub-Saharan African region, before the next vaccine campaign begins and the same barriers are encountered again.

COVID-19 is not the last disease for which Malawi will need its youth to say yes. The question this study asks and begins to answer is what it will take to make that yes more likely.

## Supporting information

## Data Availability

Data is available on request from the corresponding author

## Acknowledgements

The authors sincerely thank the 378 young people of Nyungwe, Karonga District, who gave their time and shared their perspectives with the research team. Their willingness to engage honestly with questions about a sensitive topic is the foundation on which this evidence rests. We also thank the community leaders of Nyungwe for facilitating access to the community, and the data collectors for their diligence, care, and professionalism throughout the fieldwork period.

## Author contributions

Conceptualization: LN, BCM. Data curation: LN. Formal analysis: LN, EC. Funding acquisition: N/A. Investigation: LN. Methodology: LN, BCM. Project administration: BCM. Resources: BCM. Supervision: BCM. Validation: EC, BCM. Visualization: LN, EC. Writing – original draft: LN, EC. Writing – review and editing: BCM, EC. All authors reviewed and approved the final manuscript prior to submission.

## Data availability statement

Readers requiring further information about the dataset or data collection procedures may contact the corresponding author at.

## Ethics statement

This study was conducted in full accordance with the ethical principles set out in the Declaration of Helsinki. Ethical approval was granted by the University of Livingstonia Research Ethics Committee (reference: UNILIA-REC/UGS/64/2023). Written informed consent was obtained from all participants prior to data collection. Participation was entirely voluntary; participants could withdraw at any time without penalty. All data were de-identified prior to analysis and stored securely with access restricted to the named research team.

